# Side effects of lower dose compounded semaglutide paired with a behavioral management program: A real-world retrospective study

**DOI:** 10.64898/2026.09.08.26362532

**Authors:** Erin C. Owen, Robert G. Moulder, Jessica L. Morse, M. Cole Ainsworth, Alexander Fabry, Amanda R. Merner

## Abstract

**Objective:** Evaluate differences in side effects between a low and standard dose compounded semaglutide paired with a behavioral companion in the first 16 weeks of treatment using a real-world, retrospective cohort.

**Methods:** Side effects were compared between the low dose protocol (Noom Microdose GLP-1 Rx Program; NMGP, up to 0.6mg/weekly compounded semaglutide) and Noom’s standard dose of compounded semaglutide (Noom GLP-1 Rx Program; NGP, up to 1.2mg/weekly). Users were propensity score matched on side effect rate in the first 8 weeks when dosing protocols were identical, and longitudinal side effect trajectories were compared via logistic regression. Analyses were conducted on a second cohort with additional matched baseline characteristics.

**Results:** The full sample included 7,682 NGP and 2,877 NMGP users. NMGP users had 34% lower and 29% lower relative odds of reporting GI side effects (*p*=0.001) and any side effects (*p*=0.001) by week 16, respectively (n=2,481 per group). Results of the second cohort demonstrated 38% lower odds of any side effect by week 16 among NMGP users (n=958 per group, *p*=0.019).

**Conclusions:** Lower doses of compounded semaglutide alongside a behavioral companion are associated with fewer side effects in the first 16 weeks compared to a higher dose program.

## Introduction

Obesity is a highly prevalent condition and is associated with increased morbidity and mortality (1). Glucagon-like peptide-1 receptor agonists (GLP-1 RAs) have transformed the treatment of obesity, with studies demonstrating substantial weight reduction. In randomized clinical trials (RCTs), users taking Federal Drug Administration (FDA) approved semaglutide lost an average of approximately 15% of their body weight over 68 weeks (2), and real-world studies reveal weight loss of approximately 4% at 3 months, 7% at 6 months, and 10% at 12 months among users with overweight or obesity (3).

However, the stimulation of GLP-1 receptors may elicit side effects, which can negatively impact treatment uptake, adherence, and persistence (4) as well as quality of life (5). Known GLP-1 RA side effects include gastrointestinal (GI) symptoms, thyroid carcinoma, diabetic retinopathy, and reduced muscle mass (4) as well as headache, fatigue, and kidney problems (6).

GI side effects are the most frequently reported symptoms associated with GLP-1 RA use. An estimated 15% to 45% of GLP-1 RA users report nausea, vomiting, indigestion, diarrhea, and/or constipation (7), with higher endorsement at the beginning of treatment and upon dose escalation (8). A systematic review revealed the risk of nausea for FDA approved semaglutide users was 2.95 times higher compared to placebo (7). In one RCT, GI adverse events were reported by 43% of semaglutide users, with 19.2% reporting nausea, 8.1% vomiting, and 13.7% diarrhea (7). In a clinical trial, 9% of participants discontinued oral semaglutide due to GI symptoms (9).

While gastrointestinal side effects are most prevalent, fatigue and headache have been observed among FDA approved GLP-1 RA users at nearly twice the rate of individuals taking a placebo (6) or non-GLP-1 RA antidiabetic medication (5). In Ghusn and colleagues (10), 6.3% of patients taking semaglutide reported fatigue, which may be attributable to reduced caloric intake (2). A review conducted by Filippatos and colleagues (11) noted incidence of headache with earlier GLP-1 medications (e.g., exenatide, liraglutide) ranging from 2% to 25%, and while studies are limited, 1.74 higher odds of headache compared to non-GLP-1 RA antidiabetic medications have been recently reported.

Despite the potential benefits of GLP-1 RAs, concerns about side effects may prevent individuals from initiating treatment (12,13), and side effects may contribute to early discontinuation of GLP-1 medications among users (14–17). Over 65% of adults in a community sample indicated risk of side effects as a barrier to seeking GLP-1 RA treatment (13). Among GLP-1 RA users, 6.5% of those enrolled in RCTs discontinued treatment due to an adverse event, and rates appear higher in observational studies (4).

Slowing dose escalation or remaining at a lower dose than the standard recommended therapeutic dose may improve GLP-1 RA tolerability. A dose-dependent increase in GI symptoms has been noted with FDA approved semaglutide (18–20). “A dose-dependent increase in GI AEs (mainly nausea and vomiting) and study withdrawals due to GI AEs was observed with semaglutide, which is consistent with the known side effects of the GLP-1 drug class” (21). Studies have also reported on dose-dependent increases in incidence of GI AEs and AEs contributing to discontinuation of an FDA approved dual agonist, which targets both GLP-1 and GIP receptors (19,20), with 39% reporting GI AEs in response to 5mg tirzepatide, 46% reporting GI AEs on 10mg tirzepatide, and 49% reported GI AEs on 15mg tirzepatide (20). GI symptoms observed with higher doses of semaglutide have been deemed unacceptable for continued administration by some (21). Prevalence of GI-related adverse events tends to increase with higher doses of GLP-1 RAs (20,22); however, no known dose-response studies of other side effects were identified in the literature.

Although prior studies have characterized side effect profiles among standard-dose users of FDA approved semaglutide, and a limited number have examined differences in side effects by dosage of semaglutide (e.g., (21)), few studies have leveraged a design in which participants are prescribed the same dose initially followed by a period of dose divergence or lower terminal dosages, and no identified studies have examined side effects among users taking different doses of compounded semaglutide.

Noom offers app-based behavioral weight management programs along with compounded semaglutide protocols provided under medical supervision. In the present study, users enrolled in a lower dose compounded semaglutide program (Noom’s Microdose GLP-1 Rx Program; NMGP) were compared to those enrolled in Noom’s standard dose compounded semaglutide program (Noom GLP-1 Rx Program; NGP). Titration protocols did not exceed 0.6mg/weekly in the first 8 weeks for either program. While the NMGP protocol was capped at 0.6mg/weekly, NGP users continued to titrate up to 1.2mg over the subsequent eight weeks. This approach enabled a rigorous evaluation of dose-dependent differences in side effects while accounting for baseline equivalence, as participants were matched on side effect rates observed during the initial eight-week period in addition to key demographic variables in a sensitivity analysis subsample. The primary aim of the present study was to determine whether individuals receiving a lower dose of compounded semaglutide (NMGP users) experienced fewer side effects after dose divergence compared to those receiving Noom’s standard dose of compounded semaglutide (NGP users).

## Methods

### Study Design

This retrospective study used a dataset of participants new to Noom who enrolled in either Noom Microdose GLP-1 Rx Program (NMGP) or Noom GLP-1 Rx Program (NGP) between July 21, 2025, and August 31, 2025, and were followed through December 31, 2025. This date range reflects Noom users who enrolled in NMGP at launch and were followed for the first 16 weeks. All procedures were approved by the Advarra institutional review board (Protocol Number: Pro00076674). NMGP and NGP participants were matched on sex, age, starting body mass index (BMI), region of the United States, Area Deprivation Index (ADI), and living environment (rural or urban).

### Participants

All participants were required to be US adults (18-80 years old) and enrolled in NMGP or NGP with a paid subscription after the three-week program trial period. Participants were excluded if they had: BMI < 25, active cancer, liver failure, severe heart disease, personal or family history of Multiple Endocrine Neoplasia Type 2, Type 2 diabetes, personal or family history of Medullary Thyroid Cancer, benzoyl alcohol allergy, were pregnant or nursing, or reside in a state in which Noom does not operate. Participants who switched programs during the study period were also excluded.

### Measures

#### Demographics

Gender, age, and BMI were self-reported at program onboarding. Geographic region and urbanicity were derived from residential zip codes using Rural-Urban Commuting Area (RUCA) codes. ADI, a measure of socioeconomic deprivation that combines housing, income, education, and employment indicators, was derived from Zip Code Tabulation Area (ZCTA) (23).

#### Side effects

Participants self-reported side effects weekly as part of routine engagement in the Noom application. Participants were asked, “Have you experienced any of these common side effects since your last dose?” The Noom application prompted participants to select as many as applicable of the following side effects at the time of weekly semaglutide injection: “Nausea”, “Constipation”, “Diarrhea”, “Indigestion”, “Fatigue”, “Headache”, “Other”, or “None”.

### Programs

#### NGP and NMGP

Users were evaluated for treatment by a licensed medical provider, and if appropriate, were prescribed a treatment protocol of compounded semaglutide. In both programs, users slowly titrated doses of compounded semaglutide, and, along with titration, personalized protocols and routine medical check-ins were used to minimize risks of side effects and rapid weight loss. NGP and NMGP users had access to the Noom app, which leverages behavioral psychology to promote healthy nutrition and lifestyle habits and access to a GLP-1 RA-specific Success Kit (for details see (24)).

#### Compounded Semaglutide Manufacturing and Dosing Protocols

FDA registered facilities supply raw ingredients that are tested and are potency corrected at the time of compounding. Noom’s compounding pharmacy partners operate under Section 503A of the Federal Food, Drug, and Cosmetic Act, 21 U.S.C. § 353a (25,26), under which oversight of compounding practice rests primarily with the State Boards of Pharmacy, and they must adhere to United States Pharmacopeia (USP) guidelines. The compounded semaglutide formulation consisted of semaglutide (2.5 mg/mL) and glycine, USP (5 mg/mL), in a vehicle containing sodium phosphate buffer, propylene glycol, USP (1.5% v/v), and phenol (0.6%) as a preservative, with pH adjusted using sodium hydroxide and/or hydrochloric acid as needed. Water for Injection, USP, was added quantum sufficit (q.s.) to final volume. A Certificate of Analysis provided by the compounding pharmacy shows sterility and bacterial endotoxin test results for each batch produced. The Compounding Log provided by the pharmacy shows ingredients used in each batch.

Standard dosing protocols of compounded semaglutide for both programs started at 0.2mg/week and increased to 0.6mg/week at week 8. From weeks 9 through 16, the dosing protocol called for NMGP to maintain the 0.6mg/weekly dose, whereas the NGP protocol increased to 0.85mg/weekly during weeks nine and ten and to 1.2mg/weekly from week 11 through 16. Individual titration schedules could diverge based on tolerability & weight loss speed. Distributions of the average dose logged weekly for each program were derived to examine if real-world dosing aligned with the above protocols (Figure 1).

**Figure 1.**
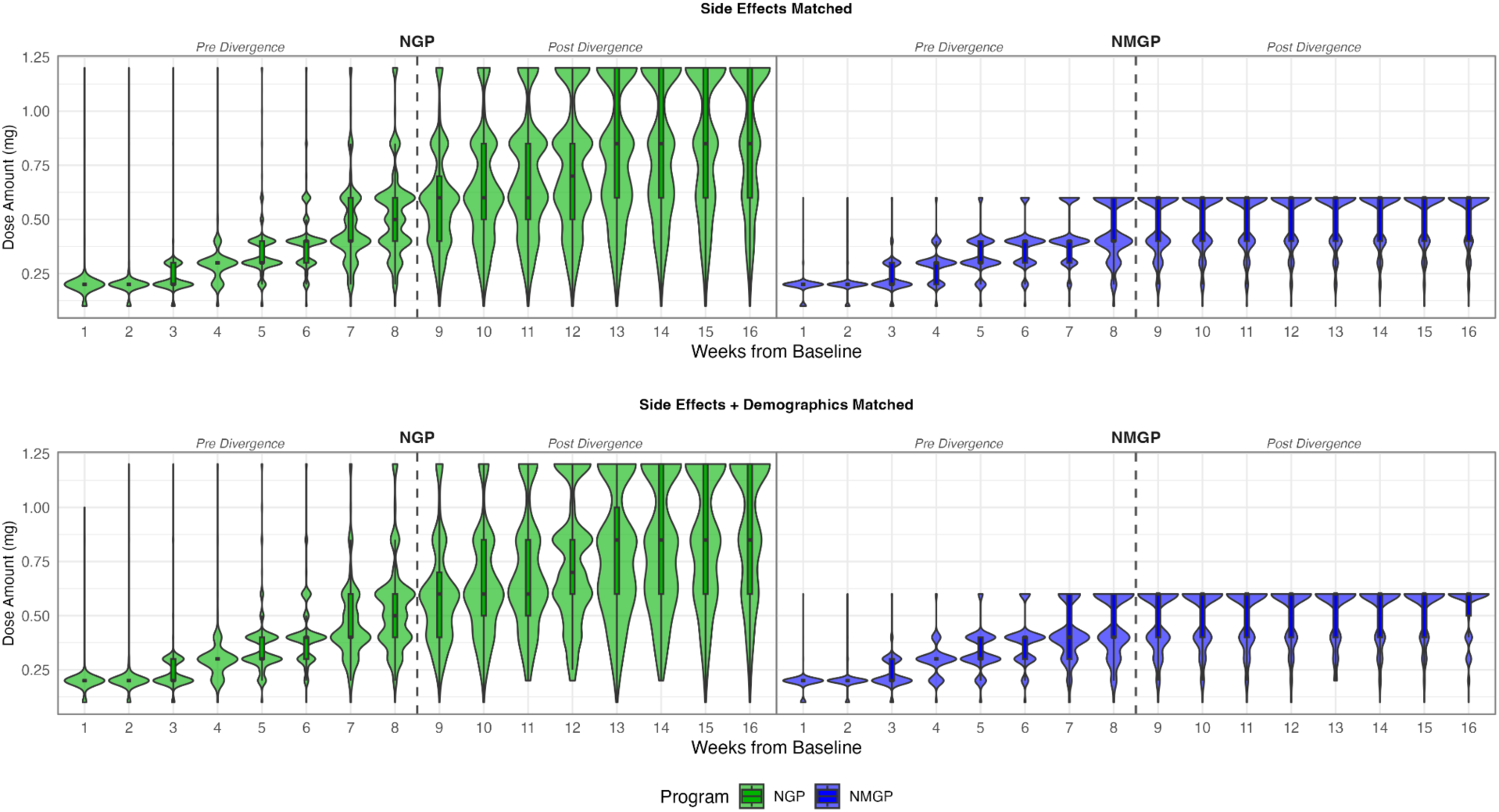
Violin plots displaying the full distribution of dose amounts each week for participants enrolled in NGP and NMGP side effect matched cohort (N=2481 per group) and for participants enrolled in NGP and NMGP side effect + demographic matched cohort (N=958 per group).

## Data Extraction and matching procedure

Data was collected from 13,316 NGP and 4,788 NMGP users and, after excluding extreme outliers (>3 standard deviations) and participants with fewer than two measurements, the full sample included 7,682 NGP and 2,877 NMGP users. Raw weekly side effect rates were calculated from this full sample. Then, generalized propensity score matching using the vecmatch R package (27) was conducted, matching NGP and NMGP users on average count of weekly reported side effects in the first eight weeks, addressing potential detection bias which may be particularly relevant in early microdose program adopters. After matching, the side effect (SE) matched cohort included 4,962 (2,481 participants per group).

To evaluate an alternative analytical approach, we conducted a second propensity score matching procedure (27). Participants were matched on average count of weekly reported side effects in the first eight weeks, gender, age, baseline BMI, US region, ADI, and rural versus urban RUCA categorization. For ADI, only 1:1 matches identified utilizing the “sociome” package in R (28) were included in the final matched cohort, resulting in a side effect + demographics matched cohort of 1,916 (SE+DEM; n=958 participants per group).

## Analytic Plan

Raw side effect rates were calculated as the percentage of users endorsing side effects. Further analyses of both matched cohorts (SE and SE+DEM) employed logistic regression to compare longitudinal side effect rate trajectories among NGP and NMGP users. Logistic regression was used to compare the proportion of NGP and NMGP users reporting any side effect after the initial eight weeks of treatment, with NMPG as the reference group. These analyses examined the presence or absence of each side effect during the second half of the study (weeks 9 through 16) to assess differences in later-stage side effect prevalence, adjusting for initial side effect profiles and potential baseline differences. Odds of experiencing side effects are reported with 95% confidence intervals [CI].

Weekly side effect rates were modeled using mixed effects logistic regression with random intercepts and slopes for each participant, nesting the weekly time points (Level 1) within individuals (Level 2) for both matched cohorts. This approach estimates changes in side effect probability over time within a person, while simultaneously accounting for the non-independence of repeated measures. Unlike the simpler logistic regression which assessed a single between-person difference in prevalence at a specific time point, the mixed effects models differentiated between individual-level variability (random effects) and the fixed effects of time and group, providing a more granular understanding of how side effect risk evolved longitudinally for NGP and NMGP users. Asymptotic confidence intervals were employed to quantify uncertainty in all estimates. These models use full information maximum likelihood (FIML) estimation, which accommodates missing at random (MAR) data by utilizing all available observations from each participant regardless of whether they completed all 16 weeks of follow-up. Participants with intermittent missing side effect reports contribute their observed data without listwise deletion or imputation (29). To visualize trends in the odds of NMGP endorsing a side effect relative to NGP over the 16-week period, the ratio of odds ratios (ROR) was plotted.

## Results

### Baseline characteristics

Baseline characteristics of the full sample (N=10,559) prior to matching, the SE matched cohort (N=4962), and the side effect + demographics matched cohort (N=1916) were very similar (Table 1). Most participants identified as female, averaged 45 years old, resided in urban areas, with few (8% - 14%) individuals residing in ZCTAs scoring as “high” deprivation.

**Table 1.** Baseline characteristics of compounded semaglutide program users: Full sample, side effect matched cohort, and side effect + demographics matched cohort.

| Baseline Demographics | Full sample |  | Side Effect Matched Cohort |  | Side Effect + Demographics Matched Cohort |  |
| --- | --- | --- | --- | --- | --- | --- |
|  | NGP | NMGP | NGP | NMGP | NGP | NMGP |
| **N** | 7682 | 2877 | 2481 | 2481 | 958 | 958 |
| **Female** | 6537 (85.1%) | 2660 (92.5%) | 2145 (86.5%) | 2303 (92.8%) | 898 (93.7%) | 898 (93.7%) |
| **Age** | 43.83 (13.25) | 44.68 (12.14) | 43.35 (13.36) | 44.86 (12.11) | 43.26 (12.32) | 43.59 (11.80) |
| **Northeast region** | 1214 (16.0%) | 531 (18.7%) | 390 (16.0%) | 436 (17.8%) | 98 (10.2%) | 89 (9.3%) |
| **Midwest region** | 1488 (19.6%) | 524 (18.4%) | 499 (20.4%) | 468 (19.1%) | 133 (13.9%) | 149 (15.6%) |
| **Southern region** | 2383 (31.4%) | 864 (30.4%) | 758 (31.0%) | 747 (30.5%) | 297 (31.0%) | 295 (30.8%) |
| **Western region** | 2493 (32.9%) | 923 (32.5%) | 797 (32.6%) | 801 (32.7%) | 430 (44.9%) | 425 (44.4%) |
| **Urban** | 6487 (85.0%) | 2489 (87.1%) | 2096 (85.0%) | 2155 (87.4%) | 850 (88.7%) | 844 (88.1%) |
| **BMI** | 32.19 (5.09) | 29.88 (4.39) | 32.14 (5.00) | 29.95 (4.44) | 30.79 (4.33) | 30.54 (4.54) |
| **Low ADI (<= 33)** | 3177 (42.7%) | 1261 (45.3%) | 1032 (43.0%) | 1085 (45.2%) | 490 (51.1%) | 509 (53.1%) |
| **Moderate ADI (> 33 & <= 66)** | 3234 (43.5%) | 1189 (42.7%) | 1031 (43.0%) | 1024 (42.7%) | 369 (38.5%) | 369 (38.5%) |
| **High ADI (> 66)** | 1026 (13.8%) | 333 (12.0%) | 336 (14.0%) | 291 (12.1%) | 99 (10.3%) | 80 (8.4%) |

*Note.* Values are presented as n (%) for categorical variables and mean (SD) for continuous variables. NGP = Noom GLP-1 Rx Program; NMGP = Noom Microdose GLP-1 Rx Program.

### Dosage curve

Figure 1 shows the full distribution of dosage amounts by week for NMGP and NGP users for the SE matched cohort and SE + demographics matched cohort, respectively. NMGP and NGP average dosages started to diverge after week 5, which differed from the titration schedule divergence after week 8. The maximum average doses for both programs were lower than the titration schedule, with NGP users’ average maximum dose at approximately 0.9mg, and NMGP users’ average maximum dose of approximately 0.5mg.

### Side Effect Rates Prior to Matching

In the full cohort prior to matching, the number and percentage of program users who reported side effects each week are reported in Table 2. At baseline (Week 0), participants had not started compounded semaglutide; however, around 15% endorsed at least one symptom. During week 1 (the first week of compounded semaglutide use), symptom endorsement increased, with similar rates of side effects observed among NGP and NMGP users, albeit a slightly higher percentage (3.1%) of NMGP users endorsed any symptom. A similar pattern of symptom reporting was observed in weeks 4 and 8, with slightly more NMGP users reporting any symptom compared to NGP users, despite identical titration schedules.

**Table 2.** Raw side effect rates with percentage of users in each compounded semaglutide program reporting presence of each symptom in the prior week within the full sample population. Doses diverged in titration protocol after week 8 (n= 7,682 NGP up to 1.2mg/weekly dose and n=2,877 NMGP up to 0.6mg/weekly dose).

| Week | Constipation |  | Diarrhea |  | Fatigue |  | Headache |  | Indigestion |  | Nausea |  | Other |  | Any GI symptoms |  | Any symptoms |  |
| --- | --- | --- | --- | --- | --- | --- | --- | --- | --- | --- | --- | --- | --- | --- | --- | --- | --- | --- |
|  | NGP | NMGP | NGP | NMGP | NGP | NMGP | NGP | NMGP | NGP | NMGP | NGP | NMGP | NGP | NMGP | NGP | NMGP | NGP | NMGP |
| 0 | 2.6% | 3.1% | 1.5% | 1.4% | 4.7% | 4.5% | 5.4% | 5.5% | 2.3% | 2.8% | 7.9% | 8.9% | 0.0% | 0.1% | 10.9% | 12.4% | 14.4% | 16.2% |
| 1 | 10.3% | 12.0% | 4.5% | 4.4% | 10.2% | 10.7% | 10.8% | 12.2% | 5.3% | 6.9% | 14.9% | 16.0% | 0.0% | 0.0% | 25.7% | 27.9% | 32.2% | 35.3% |
| 4 | 13.2% | 14.1% | 4.0% | 4.4% | 9.8% | 11.5% | 7.0% | 9.4% | 6.5% | 8.8% | 11.8% | 14.8% | 0.0% | 0.0% | 25.6% | 29.1% | 30.1% | 35.4% |
| 8 | 13.1% | 14.5% | 3.6% | 4.0% | 9.1% | 10.1% | 5.5% | 6.1% | 7.0% | 8.6% | 11.0% | 11.9% | 0.1% | 0.2% | 24.2% | 27.1% | 28.4% | 32.0% |
| 12 | 12.1% | 11.7% | 3.1% | 3.0% | 8.2% | 6.3% | 5.4% | 5.4% | 6.7% | 5.6% | 10.4% | 8.5% | 0.4% | 0.9% | 21.9% | 21.1% | 25.3% | 25.6% |
| 16 | 10.9% | 11.3% | 3.7% | 2.3% | 6.2% | 5.2% | 4.1% | 3.1% | 5.4% | 5.1% | 8.8% | 7.1% | 0.7% | 0.7% | 20.5% | 19.3% | 23.5% | 21.6% |

By week 12, this pattern shifted such that NGP users endorsed some symptoms at higher rates than NMGP users, and this largely held through week 16. Across users and timepoints, GI symptoms, particularly nausea and constipation, were endorsed at the highest rates (Table 2).

### Side effect (SE) matched cohort

Standardized mean differences (SMD) were calculated before and after propensity score matching to assess covariate balance between the NGP and NMGP groups. Prior to matching, the SMD average distance metric was 0.111, indicating modest imbalance between groups. Following propensity score matching, the SMD average distance metric was reduced to 0.003, well below the conventional threshold of 0.100, demonstrating that matching achieved excellent balance between the two groups on observed covariates. Raw side effect rates were calculated for the SE matched cohort and did not vary substantially from those of the full sample (Table S1).

#### SE matched cohort: Estimated proportion of users experiencing side effects across weeks 9-16

Results of logistic regression analyses revealed a lower proportion of NMGP users reported side effects compared to NGP users across the entire post-titration divergence period, weeks 9 through 16 (Table S2). After dosing protocols diverged, NMGP users had 0.85 [0.75, 0.97] or 15% lower odds of reporting GI side effects and 0.86 [0.76, 0.98], or 14% lower relative odds, of reporting any side effects. NMGP users had significantly reduced relative odds, 15% to 24% lower, of reporting diarrhea, fatigue, headache, indigestion, nausea, or any GI compared to NGP users (Table S2).

#### SE matched cohort: Estimated weekly percent chance of experiencing a side effect by program

While the prior analysis provides information on the proportions of individuals ever reporting at least one side effect across the weeks 9 through 16 post-titration divergence period, the occurrence of side effects within individuals varied week-by-week. Figure 2A illustrates that the weekly percent chance of reporting a side effect declined in both groups over the study period. While NMGP had a higher percent chance of reporting any side effect early in the program (34.9% versus 32.5% in week 1), the percent chance of endorsing any side effects declined from about one-third of users in both groups to 9.5% in NGP and 7.0% in NMGP by week 16 (Table 3).

**Figure 2.**
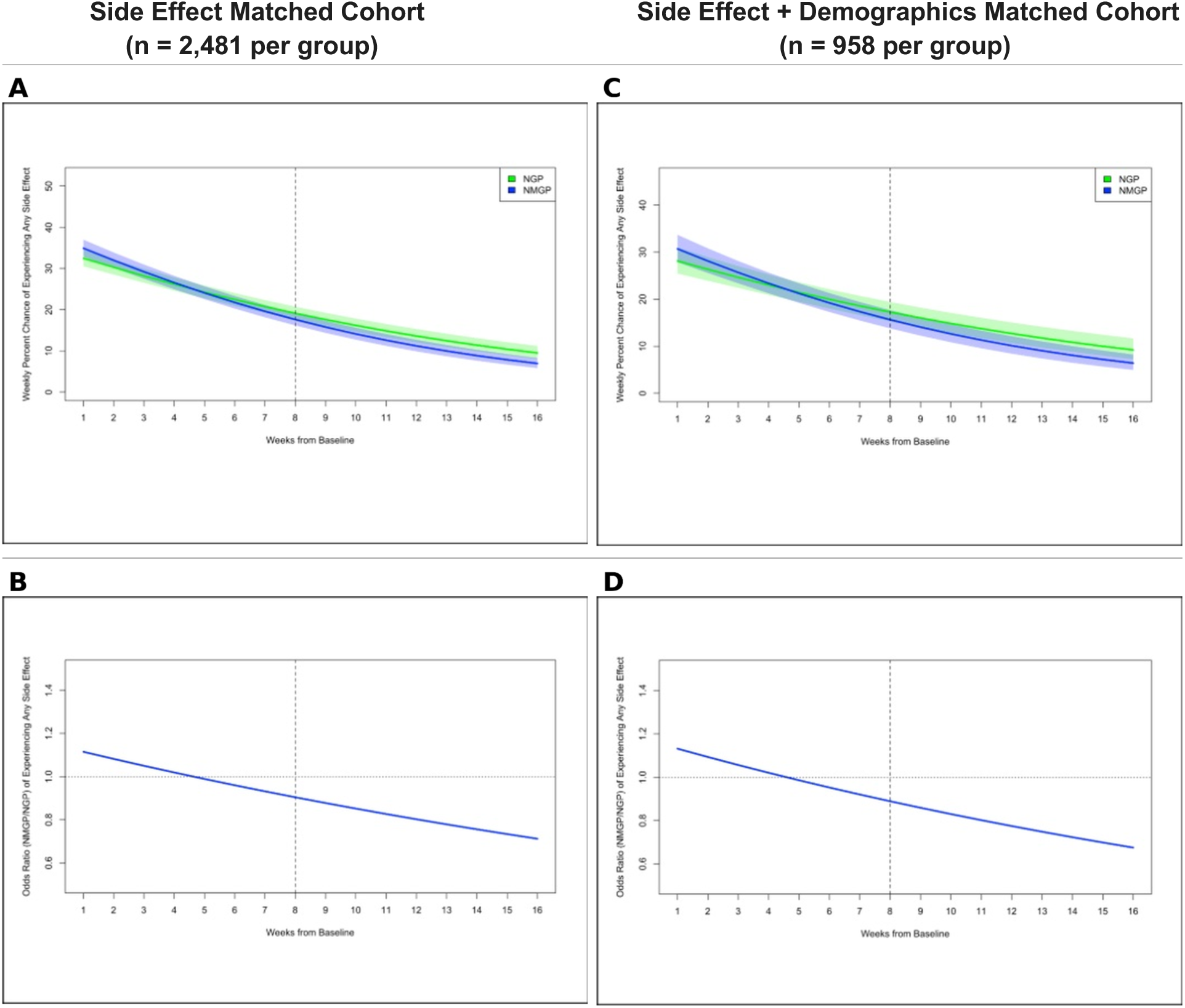
Figures 2A and 2C show the weekly percent expected chance of a person experiencing a side effect by program; titration schedule diverges after week 8. Figures 2B and 2D show the weekly ratio of odds ratio (ROR) for NMGP/NGP based on the proportion of NMGP compared to NGP users who endorsed any symptoms; titration schedule diverges after week 8. ROR = 1.0 indicates no differential effect between groups.

**Table 3.** Estimated weekly percent chance of an individual experiencing a side effect by NGP and NMGP compounded semaglutide programs in the SE matched cohort (n=2,481 per group). NMGP is the reference group for comparison.

| Constipation |  |  |  | Diarrhea |  |  |
| --- | --- | --- | --- | --- | --- | --- |
| Week | NGP<br>(%, [95% CI]) | NMGP<br>(%, [95% CI]) | Comparison | NGP<br>(%, [95% CI]) | NMGP<br>(%, [95% CI]) | Comparison |
| 1 | 4.3% [4.3%, 4.3%] | 4.4% [4.4%, 4.4%] | OR = 1.02 [1.02, 1.02],<br>$p < .001^*$ | 0.5% [0.4%, 0.8%] | 0.5% [0.4%, 0.8%] | OR = 1.01 [0.78, 1.32],<br>$p = 0.928$ |
| 4 | 3.4% [3.4%, 3.4%] | 3.5% [3.5%, 3.5%] | OR = 1.04 [1.04, 1.05],<br>$p < .001^*$ | 0.4% [0.3%, 0.5%] | 0.4% [0.3%, 0.5%] | OR = 0.94 [0.76, 1.17],<br>$p = 0.588$ |
| 8 | 2.4% [2.4%, 2.4%] | 2.6% [2.6%, 2.6%] | OR = 1.08 [1.07, 1.08],<br>$p < .001^*$ | 0.3% [0.2%, 0.4%] | 0.2% [0.2%, 0.3%] | OR = 0.85 [0.68, 1.08],<br>$p = 0.183$ |
| 12 | 1.7% [1.7%, 1.7%] | 1.9% [1.9%, 1.9%] | OR = 1.11 [1.11, 1.11],<br>$p < .001^*$ | 0.2% [0.1%, 0.3%] | 0.1% [0.1%, 0.2%] | OR = 0.77 [0.57, 1.06],<br>$p = 0.110$ |
| 16 | 1.2% [1.2%, 1.2%] | 1.4% [1.4%, 1.4%] | OR = 1.14 [1.14, 1.14],<br>$p < .001^*$ | 0.1% [0.1%, 0.2%] | 0.1% [0.0%, 0.1%] | OR = 0.70 [0.46, 1.07],<br>$p = 0.103$ |
| Fatigue |  |  |  | Headache |  |  |
| Week | NGP<br>(%, [95% CI]) | NMGP<br>(%, [95% CI]) | Comparison | NGP<br>(%, [95% CI]) | NMGP<br>(%, [95% CI]) | Comparison |
| 1 | 3.7% [3.1%, 4.5%] | 3.9% [3.2%, 4.6%] | OR = 1.03 [0.85, 1.25],<br>$p = 0.725$ | 3.1% [2.5%, 3.8%] | 3.8% [3.1%, 4.6%] | OR = 1.24 [1.02, 1.50],<br>$p = 0.028^*$ |
| 4 | 2.4% [2.0%, 2.8%] | 2.3% [1.9%, 2.7%] | OR = 0.96 [0.81, 1.14],<br>$p = 0.611$ | 1.7% [1.4%, 2.0%] | 2.0% [1.7%, 2.3%] | OR = 1.14 [0.96, 1.35],<br>$p = 0.125$ |
| 8 | 1.3% [1.1%, 1.6%] | 1.1% [0.9%, 1.4%] | OR = 0.86 [0.71, 1.05],<br>$p = 0.140$ | 0.8% [0.6%, 1.0%] | 0.8% [0.6%, 1.0%] | OR = 1.02 [0.83, 1.24],<br>$p = 0.853$ |
| 12 | 0.7% [0.5%, 0.9%] | 0.6% [0.4%, 0.7%] | OR = 0.78 [0.59, 1.01],<br>$p = 0.063$ | 0.4% [0.3%, 0.5%] | 0.3% [0.2%, 0.4%] | OR = 0.91 [0.69, 1.21],<br>$p = 0.518$ |
| 16 | 0.4% [0.3%, 0.6%] | 0.3% [0.2%, 0.4%] | OR = 0.70 [0.49, 1.00],<br>$p = 0.048^*$ | 0.2% [0.1%, 0.2%] | 0.1% [0.1%, 0.2%] | OR = 0.81 [0.56, 1.19],<br>$p = 0.293$ |
| Indigestion |  |  |  | Nausea |  |  |
| Week | NGP<br>(%, [95% CI]) | NMGP<br>(%, [95% CI]) | Comparison | NGP<br>(%, [95% CI]) | NMGP<br>(%, [95% CI]) | Comparison |
| 1 | 0.8% [0.6%, 1.1%] | 1.1% [0.8%, 1.5%] | OR = 1.40 [1.10, 1.79],<br>$p = 0.006^*$ | 7.3% [6.4%, 8.3%] | 8.2% [7.2%, 9.4%] | OR = 1.14 [0.97, 1.33],<br>$p = 0.117$ |
| 4 | 0.6% [0.4%, 0.7%] | 0.7% [0.5%, 0.9%] | OR = 1.19 [0.96, 1.47],<br>$p = 0.118$ | 5.0% [4.5%, 5.6%] | 4.9% [4.4%, 5.5%] | OR = 0.97 [0.84, 1.12],<br>$p = 0.707$ |
| 8 | 0.4% [0.3%, 0.5%] | 0.3% [0.3%, 0.5%] | OR = 0.95 [0.75, 1.20],<br>$p = 0.642$ | 3.0% [2.6%, 3.5%] | 2.4% [2.1%, 2.8%] | OR = 0.79 [0.67, 0.93],<br>$p = 0.006^*$ |
| 12 | 0.2% [0.2%, 0.3%] | 0.2% [0.1%, 0.3%] | OR = 0.75 [0.55, 1.03],<br>$p = 0.075$ | 1.8% [1.5%, 2.2%] | 1.2% [0.9%, 1.4%] | OR = 0.64 [0.51, 0.81],<br>$p < .001^*$ |
| 16 | 0.2% [0.1%, 0.2%] | 0.1% [0.1%, 0.2%] | OR = 0.60 [0.40, 0.91],<br>$p = 0.015^*$ | 1.1% [0.8%, 1.4%] | 0.6% [0.4%, 0.7%] | OR = 0.52 [0.38, 0.71],<br>$p < .001^*$ |

| Week | Any GI |  |  | Any Symptom |  |  |
| --- | --- | --- | --- | --- | --- | --- |
|  | NGP<br>(%, [95% CI]) | NMGP<br>(%, [95% CI]) | Comparison | NGP<br>(%, [95% CI]) | NMGP<br>(%, [95% CI]) | Comparison |
| 1 | 24.5%<br>[22.8%, 26.2%] | 25.8%<br>[24.1%, 27.7%] | OR = 1.08 [0.95, 1.22],<br>$p = 0.259$ | 32.5%<br>[30.5%, 34.5%] | 34.9%<br>[32.9%, 37.0%] | OR = 1.11 [0.98, 1.26],<br>$p = 0.089$ |
| 4 | 19.3%<br>[18.0%, 20.7%] | 18.9%<br>[17.6%, 20.3%] | OR = 0.97 [0.87, 1.10],<br>$p = 0.664$ | 26.2%<br>[24.6%, 27.8%] | 26.6%<br>[25.0%, 28.2%] | OR = 1.02 [0.91, 1.14],<br>$p = 0.750$ |
| 8 | 13.8%<br>[12.6%, 15.1%] | 12.0%<br>[11.0%, 13.2%] | OR = 0.85 [0.74, 0.99],<br>$p = 0.031^*$ | 19.1%<br>[17.6%, 20.7%] | 17.6%<br>[16.2%, 19.1%] | OR = 0.90 [0.79, 1.04],<br>$p = 0.154$ |
| 12 | 9.7%<br>[8.5%, 11.0%] | 7.4%<br>[6.5%, 8.5%] | OR = 0.75 [0.62, 0.91],<br>$p = 0.004^*$ | 13.6%<br>[12.1%, 15.3%] | 11.2%<br>[9.9%, 12.7%] | OR = 0.80 [0.67, 0.97],<br>$p = 0.021^*$ |
| 16 | 6.7%<br>[5.6%, 8.0%] | 4.5%<br>[3.7%, 5.5%] | OR = 0.66 [0.51, 0.85],<br>$p = 0.001^*$ | 9.5%<br>[8.0%, 11.2%] | 7.0%<br>[5.8%, 8.3%] | OR = 0.71 [0.56, 0.91],<br>$p = 0.007^*$ |

*Note.* NMGP is the reference group for all comparisons. “Other” side effects excluded due to low endorsement ( $n < 12$ ). Week 8 is shaded to signify divergence in titration schedules between NGP and NMGP after week 8; \* = $p < .05$ and 95% CI not containing 1.00.

Results of mixed-effects logistic regression analyses revealed the relative odds of experiencing any side effect by week 16 was 0.71 [0.56, 0.91], or 29% lower in NMGP compared to NGP. Although the relative odds of reporting any side effect became statistically significant three weeks after scheduled dose divergence (week 12), Figure 2B shows the ratio of odds ratios (ROR), demonstrating a progressive decline favoring lower side effects in NMGP after week 4.

The odds of reporting any GI side effect were significantly lower in NMGP users just after actual dose titration started to diverge (Week 8). At week 8, NMGP users had 0.86 [0.74, 0.99], or 14% lower odds of endorsing any GI side effect compared to NGP users. By week 16, the relative odds of reporting a GI side effect in NMGP declined to 0.66 [0.51, 0.85] or 34% lower than NGP users. The weekly percent chance of reporting an individual side effect was small (approximately 1% or lower) in both groups for headache, diarrhea, fatigue, and indigestion from weeks 9-16. Constipation was the only symptom reported in which the relative odds were persistently higher in the NMGP population and increased over time from 1.02 [1.02, 1.02] higher odds in week 1 to 1.14 [1.14, 1.14] higher odds compared to NGP in week 16. Last, NMGP had about half the odds of reporting nausea by week 16 compared to NGP (OR = 0.52 [0.38, 0.71]), although the percent weekly chance declined from 3.0% to 1.1% and 2.4% to 0.6% in NPG and NMGP, respectively.

## Side effect + demographics matched cohort

Standardized mean differences (SMD) were calculated before and after propensity score matching to assess covariate balance between the NGP and NMGP groups. Prior to matching, the SMD average distance metric was 0.178, indicating modest imbalance between groups. Following propensity score matching, the SMD average distance metric was reduced to 0.007, below the conventional threshold of 0.100, demonstrating matching achieved excellent balance between the two groups on observed covariates. Raw side effect rates were calculated for this SE + DEM cohort and did not vary substantially from those of the full sample (Table S3).

### Side effect + demographics matched cohort: Estimated proportion of users experiencing side effects across weeks 9-16

Results of logistic regression analyses comparing NGP and NMGP users across the entire post-titration divergence period in this cohort are reported in Table S4. NMGP users had significantly lower relative odds (OR = 0.81 [0.66, 0.99]) of reporting any side effect in weeks 9 through 16. (Table S4). NMGP users had significantly lower relative odds of reporting some symptoms, including 29% lower relative odds of reporting headache, 25% lower relative odds of indigestion, 28% lower relative odds of nausea, and 19% lower odds of reporting any GI symptom (Table S4).

### Side effect + demographics matched cohort: Estimated weekly percent chance of experiencing a side effect by program

The weekly percent chance of reporting any side effects declined over the study period in the SE + DEM matched cohort (Figure 2C) from 30.4% in NMGP users and 27.6% in NGP users to 6.0% and 9.3% by week 16, respectively (Table 4). By week 16, the relative odds of reporting any side effect among NMGP users were 0.62 [0.42, 0.92] or 38% lower compared to the higher dose NGP program. Figure 2D shows the ratio of odds ratios (ROR) demonstrating a progressive decline favoring lower reporting of any side effects in NMGP after week 4, consistent with the SE matched cohort (Table 4; Figure 2D).

**Table 4.**
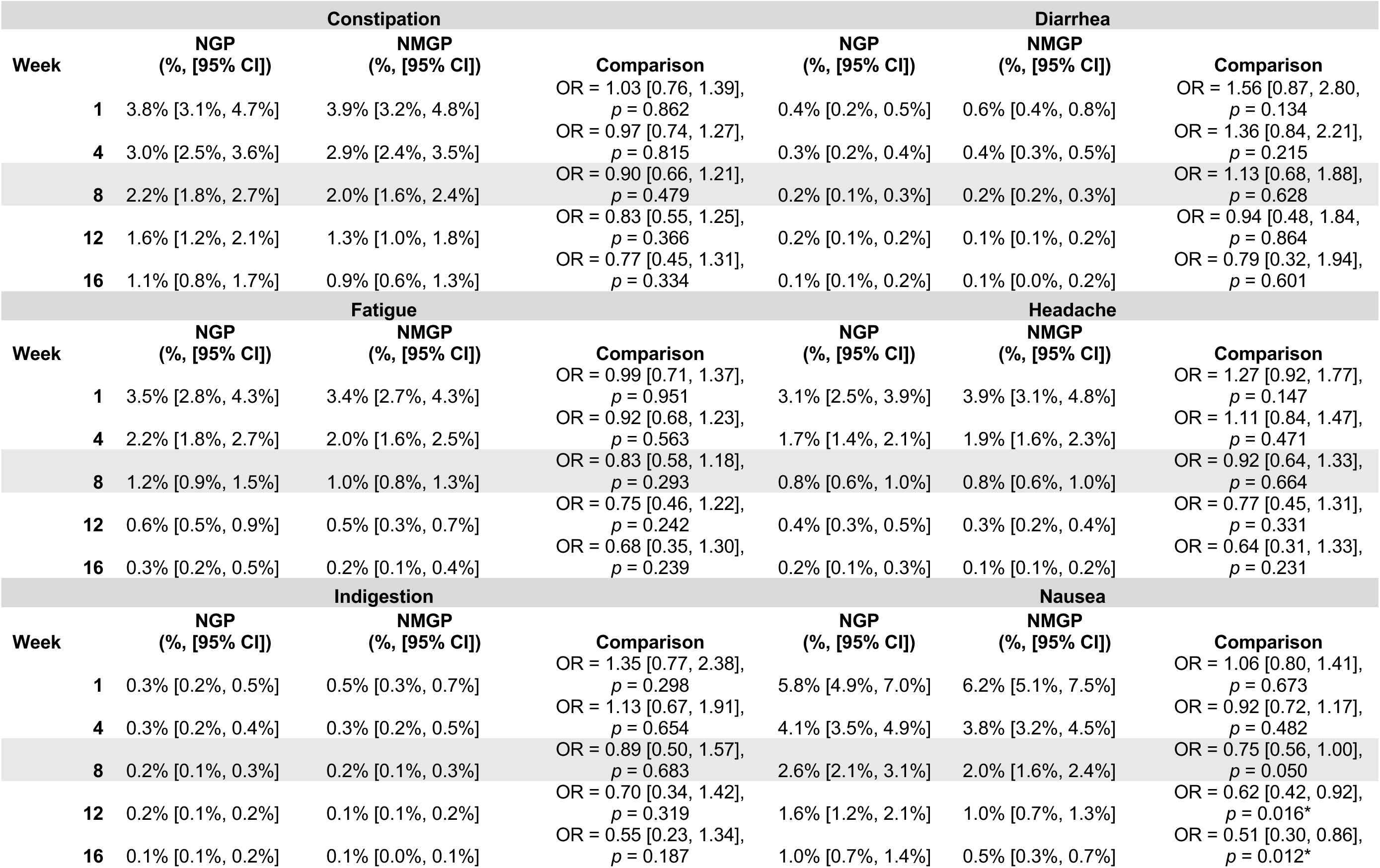
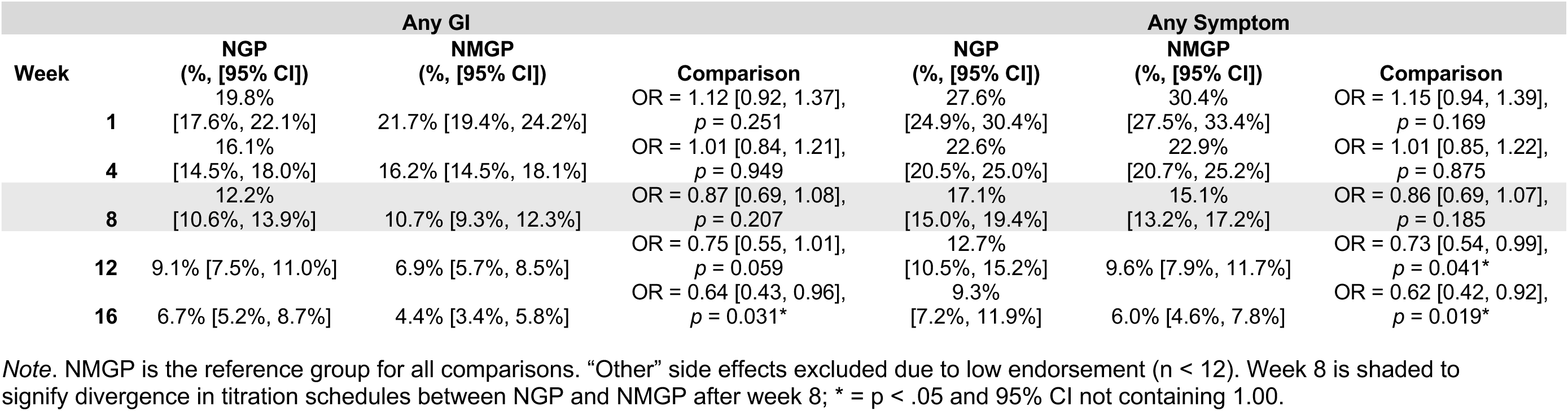
Estimated weekly percent chance of an individual experiencing a side effect by NGP and NMGP compounded semaglutide programs in side effect + demographics cohort matched on average side effect rates, gender, age, baseline BMI, urbanicity, ADI, and geography (n=958 per group). NMGP is the reference group for comparison.

## Discussion

To our knowledge, this is the first real-world evidence comparing side effects between a lower dose of compounded semaglutide and a higher dose protocol. Results suggest lower doses of compounded semaglutide are associated with improved tolerability compared to higher doses over time. After dose divergence, NMGP users had a persistent decline in side effect reporting over time. By week 16, NMGP users had 29% and 38% lower odds of reporting a side effect in the SE matched cohort and SE + DEM matched cohort, respectively, compared to the higher dose NGP users. In both matched cohorts, NMGP users also had at least one-third lower odds of reporting GI symptoms, the most frequently reported side effects associated with semaglutide, compared to NGP users by week 16. Given the challenges in medication persistence related to side effect tolerability (4,14), lower doses of compounded semaglutide may represent a promising alternative to higher dose protocols.

Analyses were conducted with a sample of users matched on the presence of early side effects, and, most rigorously, with a cohort matched on the presence of early side effects and key baseline demographic variables. The longitudinal trajectory and pattern of declining odds of side effects in NMGP users were consistent across both matched cohorts. By halfway through the post-titration divergence period, the SE + DEM matched cohort yielded findings of 25% lower relative odds of reporting any GI side effect among NMGP users compared to NGP users which was the same for the SE matched cohort. The divergence continued over time, resulting in 36% lower odds of reporting any GI side effect by week 16 in the side effect + demographics matched cohort and 34% in the SE matched cohort. A similar trend emerged for any side effect, with 27% lower odds by week 12 and 38% lower odds by week 16 among NMGP users in the SE + DEM matched cohort and 20% and 29% lower odds among NMGP users in the SE matched cohort, at weeks 12 and 16, respectively. Together, the two cohorts substantiate lower odds of side effects among NMGP users relative to users in a higher dose program (NGP), with comparable observed weight loss (24). Notably, the actual weekly percent chance of reporting any side effect after dose divergence was less than one out of 10 users by week 16 in either group (Tables 3 and 4).

Lower rates of reported side effects associated with compounded semaglutide in our study may have been due to the lower dosages prescribed to NMGP and NGP users compared to the FDA-approved maximum dosages (FDA, 2016). Ismaiel and colleagues (7) noted 43% of semaglutide users reported a GI side effect, with 19.2% reporting nausea and 13.7% reporting diarrhea. These rates are higher than the approximately 19% to 29% of users reporting any GI symptom, 7% to 16% reporting nausea, and 2% to 4.5% reporting diarrhea in the present study. Current results align more closely with Petri and colleagues (22), who found nausea reported by 5% to 15% of FDA approved semaglutide users taking a lower dose. The reporting of other side effects in this sample, including fatigue and headache, which are less frequently reported relative to GI symptoms, largely aligned with recent reports (10,11).

Side effect rates for the compounded semaglutide evaluated may be impacted by two additional factors. First, the behavioral companion prescribed alongside the compounded GLP-1 has educational content specifically targeting side effect management, including nutritional strategies. While use of a behavioral companion has been associated with improved weight loss outcomes (30), to our knowledge the impact of a companion on side effect tolerability has not been reported. Second, the compounded semaglutide administered contains other ingredients, including 5 mg of glycine, a non-essential amino acid. There exists biological plausibility that glycine may aid in side effect mitigation when combined with semaglutide. Glycine significantly enhances gastric adaptive relaxation in mice (31). Mechanistically, this may help counter the nausea and bloating related to slower gastric emptying and accommodation while taking semaglutide if it were to be substantiated in human clinical trials.

In the present study, there was substantive endorsement of symptoms by participants in both programs prior to compounded semaglutide initiation. While symptom rates increased substantially after initiation, nearly one out of six participants in the present study reported symptoms prior to dosing. Tang and colleagues (32) found GLP-1 RA use was not associated with increased risk of gastrointestinal symptoms compared to other anti-obesity medications. Future research should seek to better understand the base rate of semaglutide-associated symptoms, as the GI symptom burden attributed to semaglutide may be overattributed to drug effects.

While the present study provides insight into the improved tolerability of lower doses of compounded semaglutide over time, these doses must still provide clinically meaningful weight loss to achieve clinician endorsement. As noted above, the same cohort of NMGP users achieved 91% of the weight loss compared to the NGP population by week 16 (24). Both groups reached a >5% body weight reduction by week 8 (24). This study may be the first to demonstrate weight loss at lower doses of compounded semaglutide, using propensity score matched head-to-head comparisons of a defined lower dose arm (up to 0.6mg/weekly) versus a higher-dose arm (up to 1.2mg/weekly) within a digital health behavioral management platform to report both weight loss and side effect outcomes.

While compounding semaglutide with other ingredients has been under scrutiny (33), this is primarily due to the lack of scientific data on safety and efficacy of compounded semaglutide and the limited public disclosure of exact ingredients by some manufacturers. With full formulation transparency, the current matched cohorts provided evidence of clinically meaningful weight loss (24) and fewer side effects on a commercially available lower dose compounded semaglutide. More research is needed to understand the extent to which current findings translate to lower doses of FDA approved semaglutide when paired with a behavioral companion.

## Limitations

Given that this was a real-world retrospective cohort of all Noom users in identified programs, data on race, ethnicity, socioeconomic status (SES), comorbid disease burden and severity, as well as polypharmacy were not available. To address this potential limitation, we used two subsamples of users: one matched on the experience of early side effects and a second matched on both the experience of early side effects and key demographic variables. To address missing race, ethnicity and SES data, we relied on literature suggesting that ADI can be used as a correlate for the environmental experience of racial and ethnic disparities, although imperfect (34,35). We also excluded Noom users with Type 2 diabetes and other diseases to limit confounding by comorbid disease burden and polypharmacy. That said, residual confounding is possible without more precise measures of these variables.

The incorporation of matching on demographic variables and early side effects also partially addressed limitations introduced by the convenience sample, which may not be representative of the broader target population but is generally representative of the Noom population. The study sample showed minimal diversity in sex and geographic location, with most participants being female and residing in urban areas. This may limit generalizability, as treatment responses (e.g., side effects) may differ significantly in men and in individuals from rural geographic settings. Attrition and survivor bias are additional limitations, whereby users who experienced severe side effects may have been more likely to drop out of treatment; however, the matched design was employed to control for this in addition to FIML estimation to handle missing data.

While propensity score matching minimizes differences that may exist between program users, confounding and selection bias remains. For example, this cohort represents early adopters of commercially available and marketed “microdose” programs, which may not be representative of characteristics of consumers who wait to engage in programs with a longer market presence. Motivations for selecting a microdose program may include avoidance of side effects, which is why this study controlled for detection bias in baseline side effect reporting. This possible detection bias may explain why the NMGP cohort had higher odds of side effect reporting in the first 4 weeks of treatment. There may be other motivations for program selection that are unmeasured including, but not limited to, health literacy, health status, program cost, and prior GLP-1 experience.

This study compared side effects between program types after titration schedules diverged per clinical protocol; however, actual doses diverged earlier than the titration schedule, which complicates the interpretation of the timing of side effects. Despite earlier than anticipated divergence, the differences in reported side effects between groups continued to increase over time. Additionally, the present study did not examine side effects at specific doses. Further, not all side effects were listed, and users may have been more likely to select a listed side effect as opposed to specifying under the “other” category. Side effects were self-reported weekly and may have been subject to recall bias; however, both NGP and NMGP groups did engage in the same reporting interfaces which allows for relative comparison between groups even if absolute rates are subject to bias. Last, there was no independent clinical confirmation of self-reported adverse events.

This study was conducted by a team of researchers employed by Noom, Inc. We support and recommend external validation of our findings to strengthen confidence in these results.

## Conclusion

Little is known about side effect tolerance when combining a lower dose of compounded semaglutide with a behavioral management companion. This large, propensity score matched design allows for head-to-head comparison of reported side effects in two different programs, Noom’s GLP-1 Rx Program (NGP) and Noom’s Microdose GLP-1 Rx Program (NMGP). After titration schedules diverged, NMGP was associated with improved tolerability. In both matched cohorts, NMGP users had approximately one-third lower odds of reporting GI side effects and one-third lower odds of reporting any side effects by week 16. With reduced side effects over time, lower dose compounded semaglutide may improve medication adherence and persistence when paired with a behavioral management companion.

## Supplementary Material

***Table S1.*** Raw side effect rates in the SE matched cohort (n = 2,481 NGP and n = 2,481 NMGP).

***Table S2.*** Proportion of NGP and NMGP users reporting side effects in the SE matched cohort after titration protocol diverged (n=2,481 per group).

***Table S3.*** Raw side effect rates in the side effect + demographics matched cohort (n = 958 NGP and n = 958 NMGP).

***Table S4.*** Proportion of NGP and NMGP users reporting side effects in side effect + demographics matched cohort after titration protocol diverged (n=958 per group).

***Figure S1.*** Pre -and post-matching results.

## Clinical trial registration

N/A

## Disclosure

Authors E.C.O., R.G. M., M.C.A., and A.F. are employees at Noom, Inc. and have received salary and stock or stock options for their employment. Authors J.L.M. and A.R.M. are contractors at Noom, Inc. and have received payment for their services.

## Data Availability

All data produced in the present study are available upon reasonable request to the authors.

## Acknowledgments

The authors thank Whitney Evans for her thoughtful feedback on an earlier draft of this manuscript.

## Author Contribution

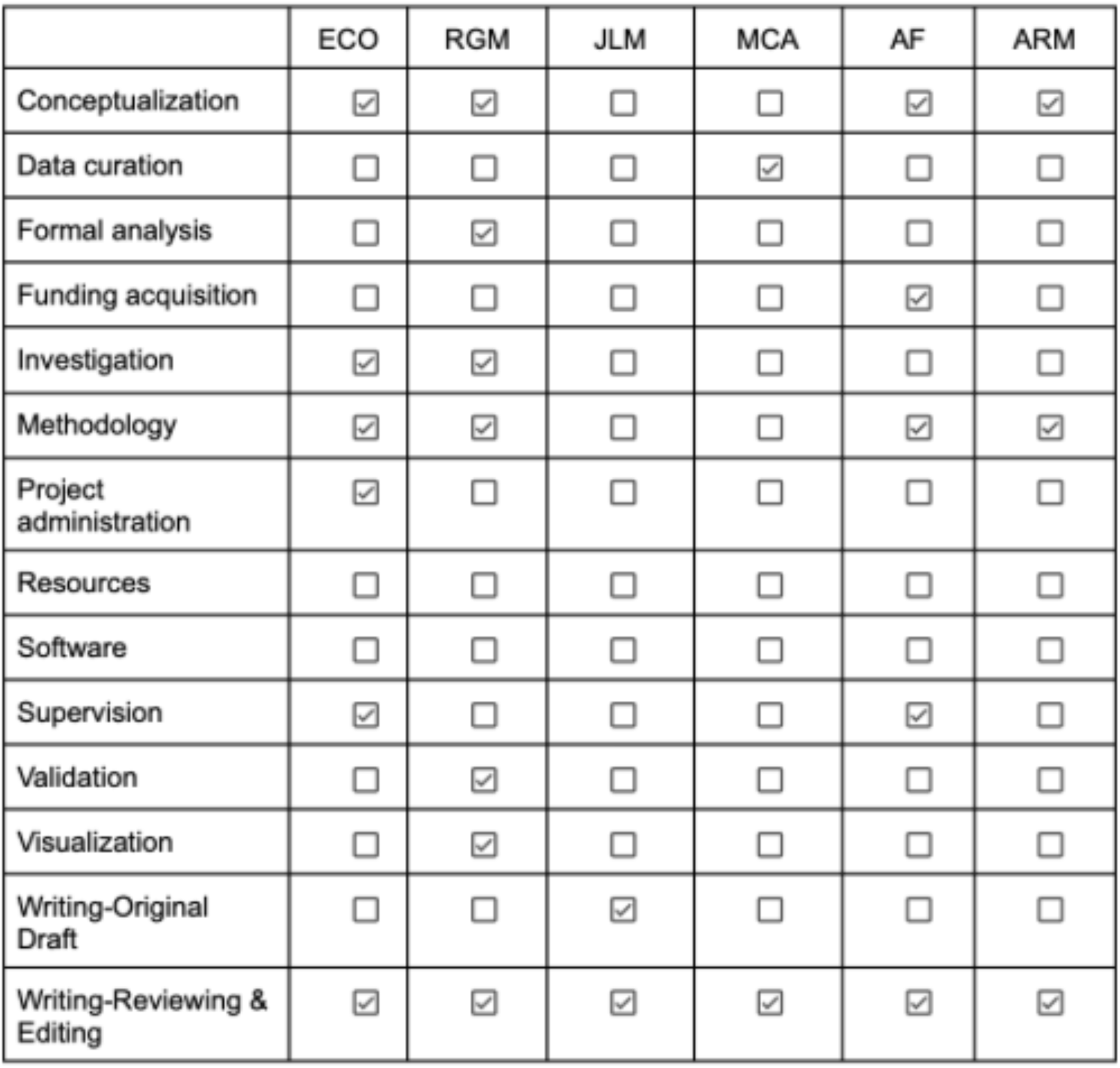

