## Supplementary materials for "Side effects of lower dose compounded semaglutide paired with a behavioral management program: A real-world retrospective study"

***Table S1.*** Raw side effect rates with percentage of users in NGP or NMGP compounded semaglutide program reporting each symptom in the side effect cohort matched only on average side effect rate in the first 8 weeks (n = 2,481 per group).

| | **Constipation** | | **Diarrhea** | | **Fatigue** | | **Headache** | | **Indigestion** | | **Nausea** | | **Other** | | **Any GI symptoms** | | **Any symptoms** | |
|---|---|---|---|---|---|---|---|---|---|---|---|---|---|---|---|---|---|---|
| **Week** | NGP | NMGP | NGP | NMGP | NGP | NMGP | NGP | NMGP | NGP | NMGP | NGP | NMGP | NGP | NMGP | NGP | NMGP | NGP | NMGP |
| **0** | 3.2% | 3.3% | 1.7% | 1.8% | 5.5% | 4.9% | 6.3% | 5.7% | 2.4% | 2.8% | 7.7% | 9.2% | 0.0% | 0.0% | 11.6% | 12.6% | 16.0% | 16.3% |
| **1** | 12.0% | 11.7% | 4.9% | 4.8% | 11.3% | 11.4% | 11.1% | 12.6% | 5.4% | 8.1% | 15.1% | 16.9% | 0.0% | 0.0% | 27.2% | 29.3% | 34.4% | 36.0% |
| **4** | 14.2% | 13.7% | 3.6% | 4.1% | 9.1% | 12.9% | 7.0% | 9.1% | 7.0% | 9.1% | 13.2% | 14.4% | 0.0% | 0.0% | 26.6% | 28.9% | 30.8% | 35.4% |
| **8** | 14.0% | 13.8% | 3.3% | 4.1% | 10.7% | 9.8% | 5.3% | 5.6% | 6.8% | 8.9% | 11.9% | 12.1% | 0.1% | 0.2% | 24.4% | 26.1% | 28.6% | 31.5% |
| **12** | 11.5% | 11.4% | 3.9% | 2.7% | 10.4% | 6.8% | 6.9% | 6.7% | 5.5% | 5.2% | 10.9% | 9.2% | 0.1% | 0.9% | 21.9% | 21.8% | 26.3% | 26.5% |
| **16** | 11.7% | 10.9% | 4.3% | 2.8% | 5.7% | 6.3% | 3.6% | 3.5% | 5.5% | 5.2% | 11.7% | 7.0% | 0.2% | 0.7% | 23.4% | 18.8% | 26.4% | 21.4% |

***Table S2.*** Proportion of NGP and NMGP users reporting side effects after week 8 in the SE matched cohort after titration protocol diverged (n=2,481 per group).

| **Side Effect** | **NGP (%, [95% CI])** | **NMGP (%, [95% CI])** | **Odds Ratio (OR [95% CI]) for NMGP side effects compared to NGP** |
|---|---|---|---|
| Constipation | 27.7% [25.8%, 29.8%] | 26.2% [24.2%, 28.2%] | OR = 0.92 [0.80, 1.07] *p* = 0.275 |
| Diarrhea | 11.9% [10.5%, 13.4%] | 9.3% [8.1%, 10.7%] | OR = 0.76 [0.62, 0.93], *p* = 0.009* |
| Fatigue | 21.9% [20.1%, 23.8%] | 18.6% [16.9%, 20.4%] | OR = 0.81 [0.70, 0.95], *p* = 0.011* |
| Headache | 16.3% [14.7%, 18.0%] | 13.7% [12.2%, 15.3%] | OR = 0.82 [0.68, 0.98], *p* = 0.026* |
| Indigestion | 17.3% [15.7%, 19.1%] | 14.4% [12.9%, 16.0%] | OR = 0.80 [0.67, 0.95], *p* = 0.013* |
| Nausea | 27.0% [25.1%, 29.1%] | 22.0% [20.2%, 24.0%] | OR = 0.76 [0.66, 0.88], *p* < .001* |
| Other | 2.7% [2.1%, 3.6%] | 2.8% [2.1%, 3.6%] | OR = 1.01 [0.68, 1.48], *p* = 0.976 |
| Any GI side effect | 46.4% [44.2%, 48.7%] | 42.5% [40.3%, 44.7%] | OR = 0.85 [0.75, 0.97], *p* = 0.015* |
| Any side effect | 51.9% [49.7%, 54.1%] | 48.2% [45.9%, 50.4%] | OR = 0.86 [0.76, 0.98], *p* = 0.019* |

***Table S3.*** Raw side effect rates with percentage of users in NGP or NMGP compounded semaglutide program reporting each symptom in the side effects + demographics cohort matched on average side effect rate in the first 8 weeks, gender, age, BMI, ADI, urbanicity, and geography (n = 958 per group).

| | **Constipation** | | **Diarrhea** | | **Fatigue** | | **Headache** | | **Indigestion** | | **Nausea** | | **Other** | | **Any GI symptoms** | | **Any symptoms** | |
|---|---|---|---|---|---|---|---|---|---|---|---|---|---|---|---|---|---|---|
| **Week** | NGP | NMGP | NGP | NMGP | NGP | NMGP | NGP | NMGP | NGP | NMGP | NGP | NMGP | NGP | NMGP | NGP | NMGP | NGP | NMGP |
| **0** | 2.9% | 2.9% | 1.0% | 1.4% | 6.7% | 3.9% | 7.3% | 5.6% | 2.3% | 2.2% | 8.5% | 7.8% | 0.0% | 0.0% | 11.0% | 11.1% | 15.6% | 14.8% |
| **1** | 12.1% | 9.3% | 4.1% | 4.1% | 12.6% | 9.1% | 12.5% | 10.7% | 5.3% | 5.8% | 15.9% | 14.2% | 0.0% | 0.0% | 27.9% | 24.5% | 35.7% | 31.4% |
| **4** | 12.6% | 12.3% | 3.3% | 3.5% | 9.9% | 11.0% | 7.6% | 9.2% | 6.4% | 7.7% | 11.9% | 13.2% | 0.0% | 0.0% | 26.3% | 25.9% | 31.3% | 32.7% |
| **8** | 13.0% | 13.2% | 2.4% | 3.8% | 10.2% | 8.4% | 6.1% | 4.4% | 7.3% | 8.6% | 11.7% | 10.9% | 0.0% | 0.0% | 24.7% | 25.3% | 29.8% | 29.2% |
| **12** | 11.8% | 10.6% | 1.9% | 3.2% | 9.7% | 5.7% | 6.4% | 5.8% | 8.3% | 6.2% | 9.0% | 8.3% | 0.7% | 1.1% | 21.6% | 21.1% | 25.9% | 25.3% |
| **16** | 12.8% | 10.8% | 2.4% | 2.2% | 7.3% | 5.1% | 5.0% | 3.2% | 6.5% | 4.6% | 8.9% | 6.2% | 0.5% | 0.5% | 22.3% | 17.0% | 26.4% | 19.5% |

***Table S4.*** Proportion of NGP and NMGP users reporting side effects after week 8 in the side effect + demographics cohort matched on average side effect rates, gender, age, baseline BMI, urbanicity, and geography after titration protocol diverged (n=958 per group).

| **Side Effect** | **NGP (%, [95% CI])** | **NMGP (%, [95% CI])** | **Odds Ratio (OR [95% CI]) Side Effects of NMGP compared to NGP** |
|---|---|---|---|
| Constipation | 27.1% [24.1%, 30.3%] | 24.8% [21.8%, 28.0%] | OR = 0.89 [0.71, 1.11], *p* = 0.304 |
| Diarrhea | 10.2% [8.3%, 12.6%] | 8.8% [7.0%, 11.1%] | OR = 0.85 [0.60, 1.20], *p* = 0350 |
| Fatigue | 20.7% [18.0%, 23.7%] | 17.8% [15.2%, 20.7%] | OR = 0.83 [0.64, 1.07], *p* = 0.145 |
| Headache | 17.1% [14.6%, 19.9%] | 12.8% [10.6%, 15.3%] | OR = 0.71 [0.53,0.94], *p* = 0.018 |
| Indigestion | 17.2% [14.7%, 20.1%] | 13.6% [11.3%, 16.2%] | OR = 0.75 [0.57, 1.00], *p* = 0.048* |
| Nausea | 25.8% [22.8%, 29.0%] | 19.9% [17.2%, 22.9%] | OR = 0.72 [0.56, 0.91], *p* = 0.006* |
| Other | 1.6% [0.9%, 2.7%] | 2.1% [1.3%, 3.4%] | OR = 1.36 [0.64, 2.90], *p* = 0.421 |
| Any GI side effect | 44.9% [41.5%, 48.5%] | 39.8% [36.4%, 43.3%] | OR = 0.81 [0.66, 0.99], *p* = 0.041* |
| Any side effect | 50.3% [46.7%, 53.8%] | 45.1% [41.6%, 48.6%] | OR = 0.81 [0.66, 0.99], *p* = 0.042* |

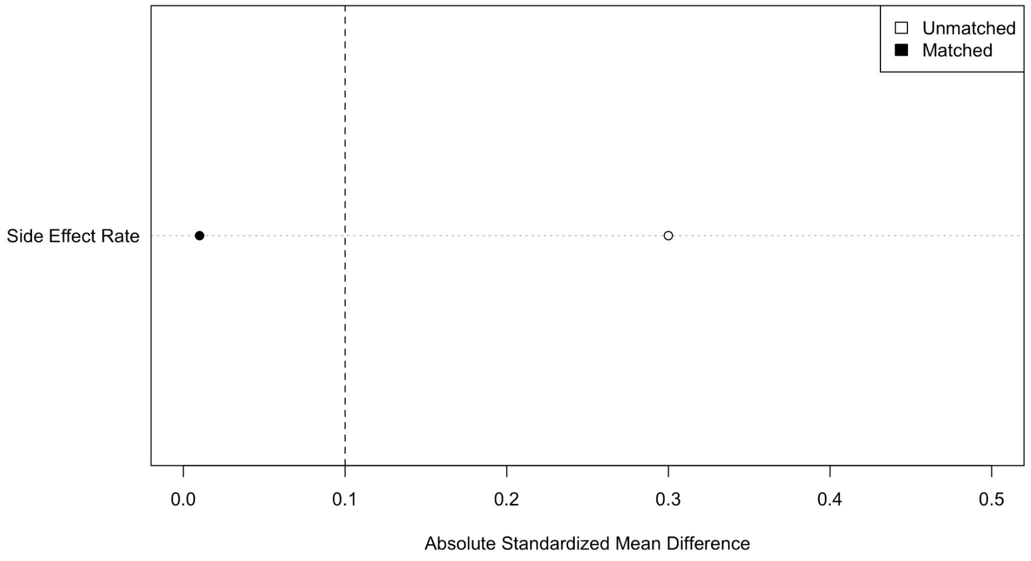


***Figure S1.*** Pre and post matching results for the side effect matched cohort. Post matching data shows acceptable matching on all matching variables.

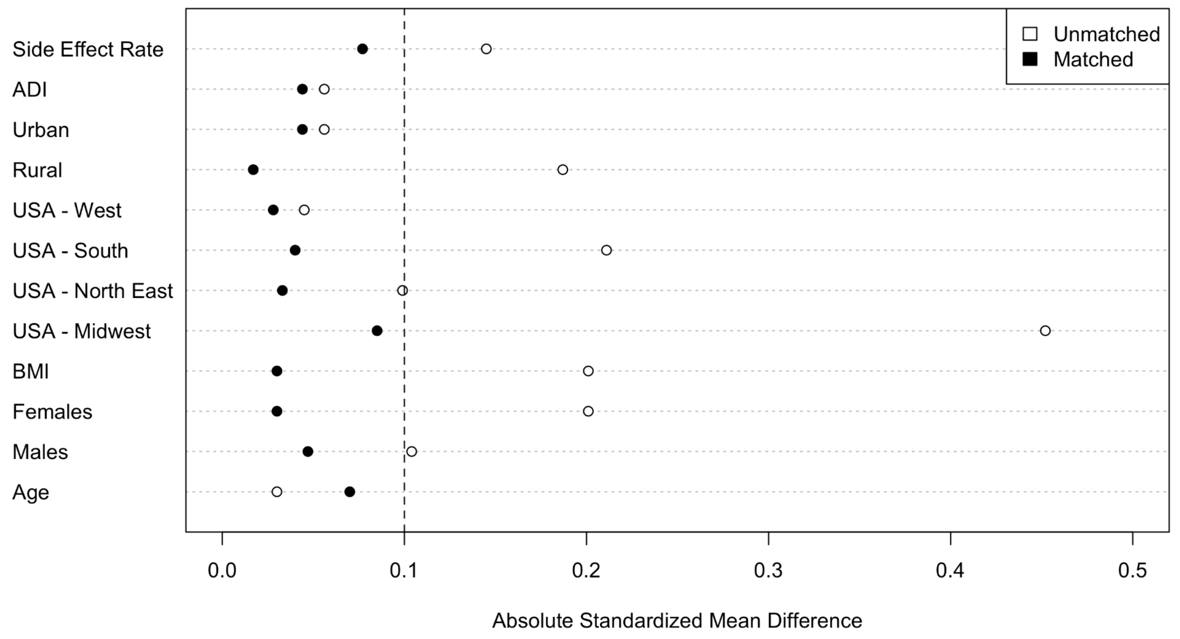


***Figure S2.*** Pre and post matching results for the side effect + demographics matched cohort. Post matching data shows acceptable matching on all matching variables.
